# Post discharge persistent symptoms after COVID-19 in rheumatic and musculoskeletal diseases

**DOI:** 10.1101/2021.03.08.21253120

**Authors:** Leticia Leon, Ines Perez-Sancristobal, Alfredo Madrid Garcia, Leticia Lopez Pedraza, Jose Ignacio Colomer, Sergio Lerma Lara, Pia Lois Bermejo, Arkaitz Mucientes, Luis Rodriguez-Rodriguez, Benjamin Fernandez-Gutierrez, Lydia Abasolo

## Abstract

**OBJECTIVES:** To describe persistent symptoms and consequences in Rheumatic and Musculoskeletal diseases (RMD) discharged from the hospital after Covid-19; to assess the roll of autoimmune rheumatic diseases (ARD) compared to and non-autoimmune rheumatic and musculoskeletal diseases (NARD) on persistent symptoms and consequences.

**METHODS:** We performed a cross-sectional observational study. RMD attended at a rheumatology outpatient clinic in Madrid with Covid-19 that required hospital admission were included. The main outcomes were persistence of symptoms and health consequences related to Covid19 after discharge. Independent variable was RMD group (ARD and NARD) and covariates were sociodemographic, clinical, and treatments. We ran a multivariable logistic regression model to assess the risk by RMD group on main outcomes.

**RESULTS:** We included 105 patients and 51.5% had ARD. 68.57% reported at least one persistent symptom. The most frequent were dyspnea, fatigue, and chest pain. 31 patients had consequences. Lung damage was found in 11.4% of the patients, 18% had blood test abnormalities (10% lymphopenia), two died, one developed central retinal vein occlusion and one patient developed optic neuritis. 11 patients required readmission due to Covid-19 problems (16.7% ARD vs 3.9% NARD; p=0.053). No statistically significant differences by RMD groups were found in the final models.

**CONCLUSION:** Many RMD patients have persistent symptoms, similar to other populations. This study also highlights that lung damage is the most frequent consequence. ARD compared to NARD does not seem to differ in terms of persistent symptoms or consequences, although ARD might have a greater number of readmissions due to Covid-19.

## Introduction

With Covid-19 pandemic continuing unabated, the global community research is focusing in acute treatment and Covid-19 vaccines strategy. The current pandemic is having a particular effect and severity in patients with advanced age and comorbidities, mainly diabetes, hypertension, ischemic heart disease, obesity and respiratory diseases [(1)(2)(3)(4)(5)]. In this sense, having an autoimmune systemic condition could be considered another risk factor for severity since these patients had a higher risk of hospital admission related to COVID-19, as we have recently published [(6)].

Previous coronavirus outbreaks have been associated with deterioration in lung function, muscle weakness, pain, fatigue, depression, anxiety, and reduced quality of life after the acute phase [(7)]. Information available in post-acute Covid-19 is scarce [(8)(9)(10) (11)(12)], but It is expected that Covid-19 would have an important impact on the state of physical, cognitive, mental and social health.[(13)(14)(15)]. In a cross-sectional study describing persistence of symptoms after hospital discharge due to Covid-19 (12), patients were evaluated an average of 60.3 days after the first symptom of Covid-19. Only 12.6% were completely free of Covid-19 symptoms, while 32% had 1 or 2 symptoms and 55% had 3 or more, particularly fatigue and dyspnea.

In Rheumatic and Musculoskeletal diseases (RMD) patients who had overcome Covid-19 disease, risk factors and outcomes of the acute phase have been described [(16)(17)(18)], but there is a lack of information after this period. Considering that most of them are chronic patients with a complex management, it would be worthy to evaluate their clinical course, the consequences and the care received after Covid-19 acute phase.

Thus, our purpose is to assess persistent symptoms and the consequences in RMD patients who were discharged from the hospital after Covid-19 and to evaluate if there are differences in them among RMD groups. We want also to describe the health care received after the acute phase.

## Methods

The setting was a tertiary hospital of the National Health System of the Community of Madrid, Spain, the Hospital Clínico San Carlos (HCSC), covering a catchment area of 350,000 people. A cross-sectional observational study was performed, including patients from the 1^st^ of March 2020, (when our health area had the first hospital admission related to Covid-19) to the 30^th^ of May 2020. All patients being attended at the rheumatology outpatient clinic of our centre during the study period, were preselected. We included patients: a) aged >18 years old; b) with medical diagnosis of rheumatic and musculoskeletal diseases using specific International Classification of Diseases, Tenth Revision (ICD-10) c) with Covid-19 disease assessed by medical diagnosis or SARS-CoV-2 PCR positive diagnostic test that required hospital admission during the inclusion period. Follow-up was from the time of discharge until October 30th. Patients who died during hospital admission were excluded (Figure 1).

**Figure 1.**
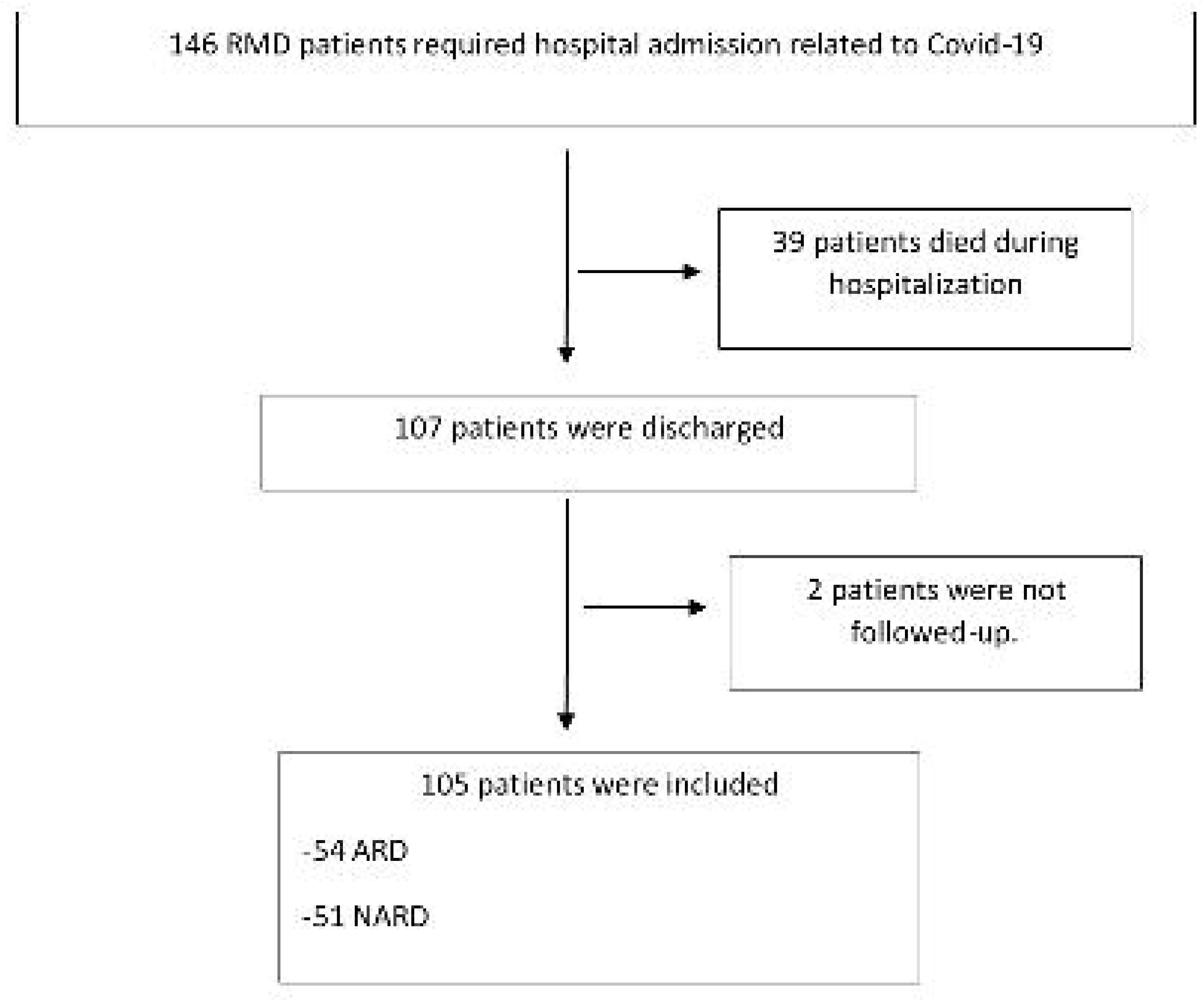
Patient’s flow chart until inclusion

The study was conducted in accordance with the Declaration of Helsinki and the principles of Good Clinical Practice and was approved by the HCSC Institutional Ethics Committee (approval number 20/268-E-BS).

Our primary outcomes were the persistence of symptoms (all new symptoms self-reported by the patient, which persists after discharge) and development of health consequences. It was considered as a new condition resulting from the Covid-19 disease reported in the clinical record, or alterations in diagnostic tests or diagnostic images that persist after the resolution of acute infection. We included death, lung damage with or without oxygen necessities, analytical abnormalities (lymphocytes, platelets, creatinine, liver markers), thrombotic event and others related to Covid-19. We registered also readmission and physical rehabilitation both related to Covid-19.

The co-variables recorded were: 1) Sociodemographic baseline characteristics. 2) RMD groups as described in table 1: a) autoimmune rheumatic diseases (ARD) including: a1) systemic autoimmune conditions (SAC); and a2) chronic inflammatory arthritis (CIA); b) Non autoimmune rheumatic and musculoskeletal diseases (NARD). 3) Chronic therapy used prior admission: glucocorticoids, NSAIDs, conventional synthetic disease modifying antirheumatic drugs (csDMARDs); targeted synthetic or biologic DMARDs (ts/bDMARDs) including: a) anti-TNF alfa; other biologics: anti-IL6; rituximab (RTX); abatacept; anti-IL17/23 or Jakinibs. 4) Baseline comorbid conditions, as described in table 1. 5) Clinical, laboratory and therapeutic data during admission. 6) Clinical care after discharge (face to face, phone calls) and physician in charge. Patient sociodemographic, clinical, laboratory and therapeutic rheumatic data were obtained through the health Clinical Record from rheumatology service of HCSC. Admission and post discharge data were collected from the information systems of the HCSC and Primary Health Care. All patients’ information including comorbidities, clinical, laboratory, chest x-rays or scans, treatment and main outcomes were checked and evaluated by two rheumatologists from the research team.

**Table 1.**
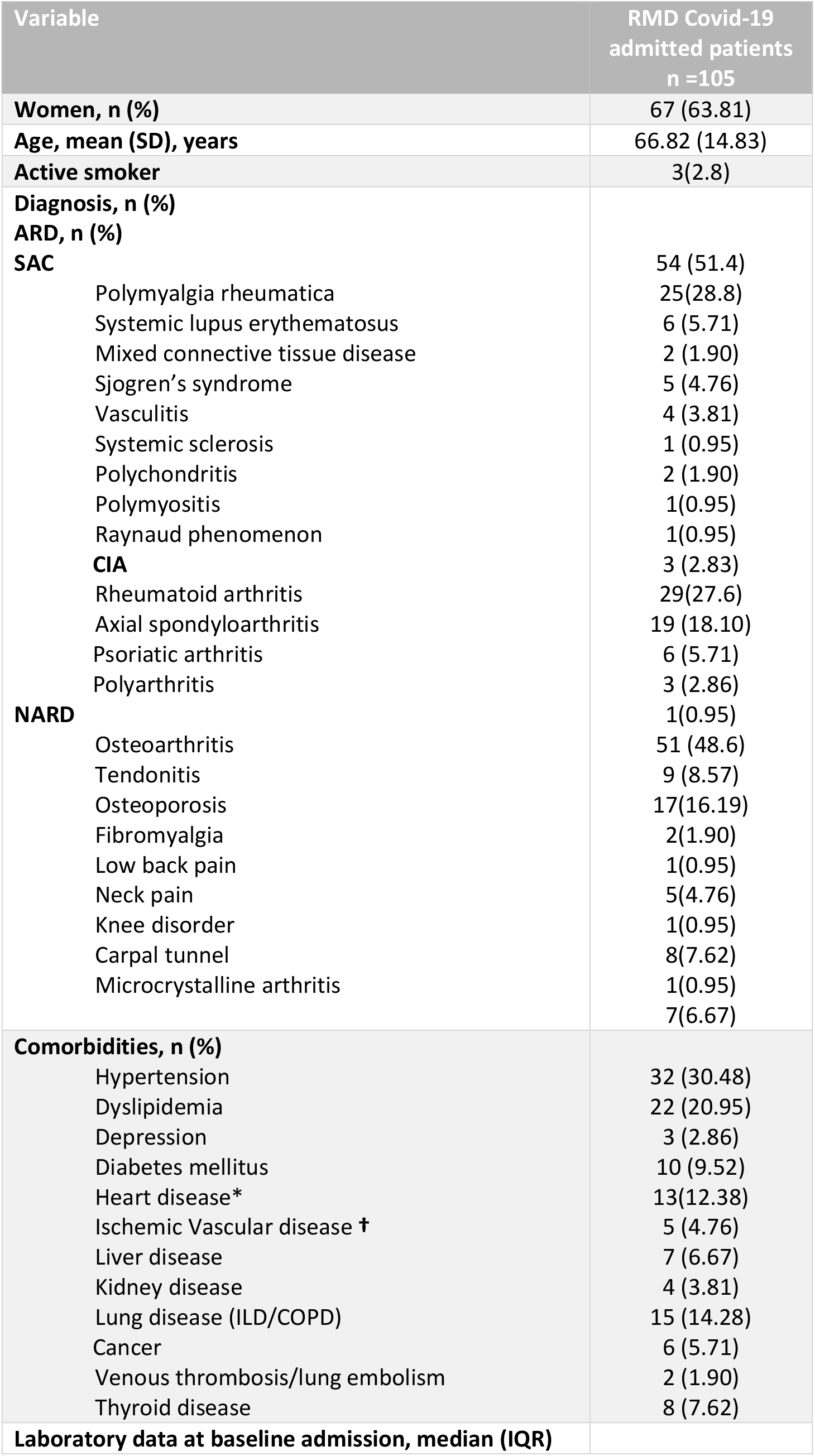
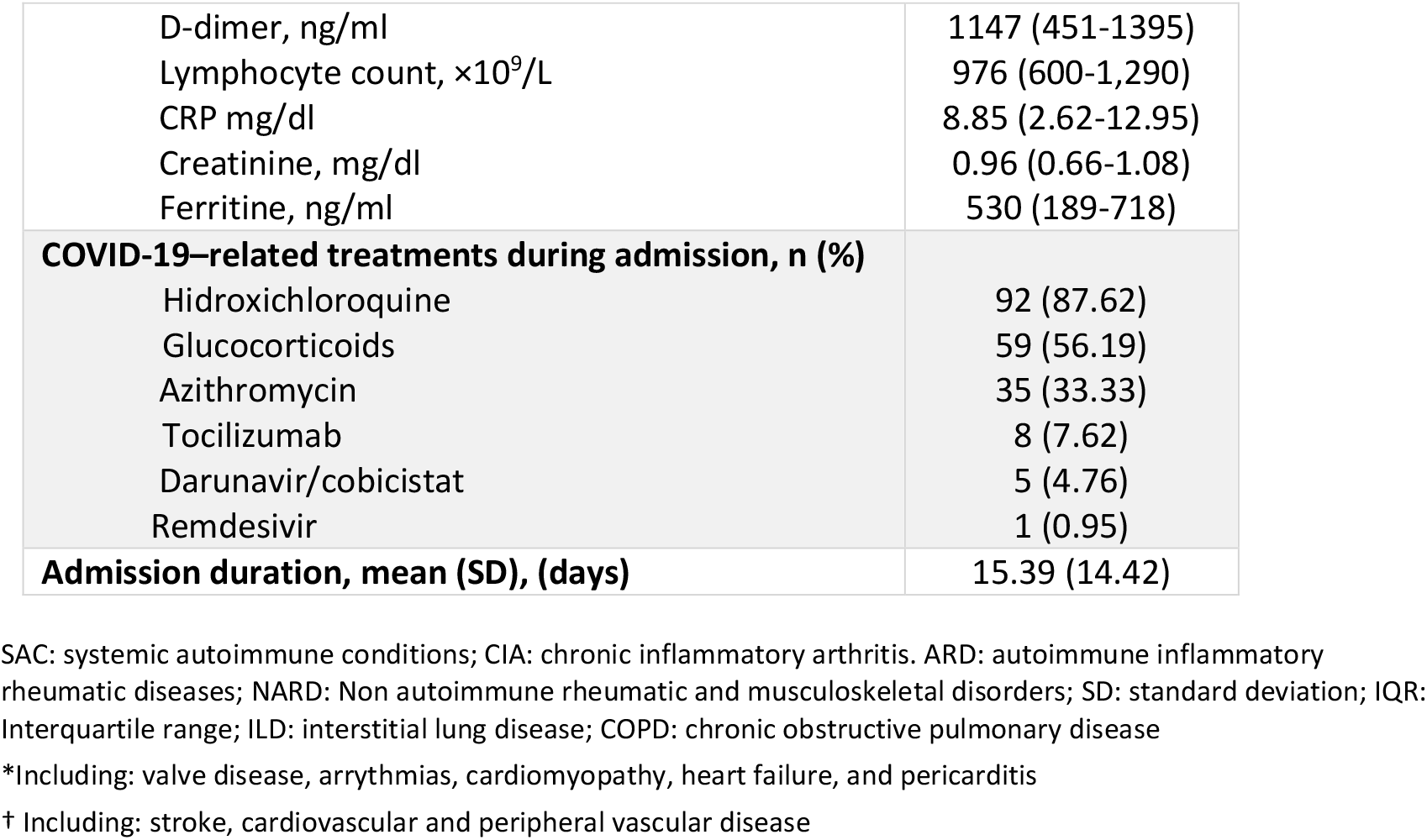
Baseline sociodemographic and clinical characteristics of RMD patients

| Variable | RMD Covid-19<br>admitted patients<br>n =105 |
| --- | --- |
| **Women, n (%)** | 67 (63.81) |
| **Age, mean (SD), years** | 66.82 (14.83) |
| **Active smoker** | 3(2.8) |
| **Diagnosis, n (%)** |  |
| **ARD, n (%)** |  |
| **SAC** | 54 (51.4) |
| Polymyalgia rheumatica | 25(28.8) |
| Systemic lupus erythematosus | 6 (5.71) |
| Mixed connective tissue disease | 2 (1.90) |
| Sjogren's syndrome | 5 (4.76) |
| Vasculitis | 4 (3.81) |
| Systemic sclerosis | 1 (0.95) |
| Polychondritis | 2 (1.90) |
| Polymyositis | 1(0.95) |
| Raynaud phenomenon | 1(0.95) |
| **CIA** | 3 (2.83) |
| Rheumatoid arthritis | 29(27.6) |
| Axial spondyloarthritis | 19 (18.10) |
| Psoriatic arthritis | 6 (5.71) |
| Polyarthritis | 3 (2.86) |
| **NARD** | 1(0.95) |
| Osteoarthritis | 51 (48.6) |
| Tendonitis | 9 (8.57) |
| Osteoporosis | 17(16.19) |
| Fibromyalgia | 2(1.90) |
| Low back pain | 1(0.95) |
| Neck pain | 5(4.76) |
| Knee disorder | 1(0.95) |
| Carpal tunnel | 8(7.62) |
| Microcrystalline arthritis | 1(0.95) |
|  | 7(6.67) |
| **Comorbidities, n (%)** |  |
| Hypertension | 32 (30.48) |
| Dyslipidemia | 22 (20.95) |
| Depression | 3 (2.86) |
| Diabetes mellitus | 10 (9.52) |
| Heart disease* | 13(12.38) |
| Ischemic Vascular disease † | 5 (4.76) |
| Liver disease | 7 (6.67) |
| Kidney disease | 4 (3.81) |
| Lung disease (ILD/COPD) | 15 (14.28) |
| Cancer | 6 (5.71) |
| Venous thrombosis/lung embolism | 2 (1.90) |
| Thyroid disease | 8 (7.62) |
| **Laboratory data at baseline admission, median (IQR)** |  |
| D-dimer, ng/ml | 1147 (451-1395) |
| Lymphocyte count, $\times 10^9/L$ | 976 (600-1,290) |
| CRP mg/dl | 8.85 (2.62-12.95) |
| Creatinine, mg/dl | 0.96 (0.66-1.08) |
| Ferritine, ng/ml | 530 (189-718) |
| **COVID-19–related treatments during admission, n (%)** |  |
| Hidroxichloroquine | 92 (87.62) |
| Glucocorticoids | 59 (56.19) |
| Azithromycin | 35 (33.33) |
| Tocilizumab | 8 (7.62) |
| Darunavir/cobicistat | 5 (4.76) |
| Remdesivir | 1 (0.95) |
| **Admission duration, mean (SD), (days)** | 15.39 (14.42) |

SAC: systemic autoimmune conditions; CIA: chronic inflammatory arthritis. ARD: autoimmune inflammatory rheumatic diseases; NARD: Non autoimmune rheumatic and musculoskeletal disorders; SD: standard deviation; IQR: Interquartile range; ILD: interstitial lung disease; COPD: chronic obstructive pulmonary disease
\*Including: valve disease, arrhythmias, cardiomyopathy, heart failure, and pericarditis
† Including: stroke, cardiovascular and peripheral vascular disease

### Statistical Analysis

Patient characteristics were expressed as mean and standard deviation or median and interquartile range for continuous variables; categorical variables are expressed as percentages. For the comparison of persistent symptoms and consequences between ARD and NARD, we used the Mann-Whitney U test, χ2 test, or Fisher’s exact test where appropriate. Multivariable logistic regression models (adjusted for age, sex, and comorbidity) were run to examine the possible effect of RMD group on outcome (persistent symptoms or consequences due to Covid-19). The model also included other variables with a p<0.2 from the univariate regression analysis. The results were expressed as the odds ratio (OR) with its respective 95% confidence interval.

All tests were two-sided, and a p value less than 0.05 was considered statistically significant.

## Results

A total of 146 RMD patients required hospital admission during the study period and 107 patients survived and were discharged, two of them were excluded (no information in data sources). Finally 105 patients were included in the study (Figure 1) and the follow-up ranged from 4 to 7 months.

Sociodemographic and clinical characteristic of patients are shown in Table 1. Two thirds of the patients were women, with a mean age of 66.82 (29.98-91.32). The mean duration of RMD was 8.9 (8.2) years and the mean duration of admission due to Covid-19 was 15.38 (14.42) days. 51.43% of patients had ARD, being rheumatoid arthritis the main diagnosis. From those with NARD, tendonitis and osteoarthritis were the most prevalent. Interestingly, 46.15% of the patients had at least one comorbid condition. Regarding chronic therapy prior admission, 32% of the patients were on glucocorticoids and 19% were on NSAIDs. Specifically in ARD patients, the following therapies were used: 8(14.8%) AM, 27(50%) MTX, 5(9.25%) AZA, 4(7.4%) LEF, 3(5.5%) SSZ and 7(12.9%) bDMARDs (RTX=3; tocilizumab=1 and TNF-alfa=3).

During the admission 92.5% of the patients had pneumonia and 41% had complications related to Covid-19 (sepsis, thrombotic, ischemic, heart, and renal complications), and 40% patients had lymphopenia (lymphocyte count <0·8×10^9^ per L). The therapy used was mainly AM and glucocorticoids.

After discharge, all the patients were followed up by at least one physician; 80 (85%) by their primary care physician (phone calls) and 45 (42%) by a specialist (face to face), mainly Pneumologist (n=22), and Internal medicine (n=9). Besides, 17 ARD patients were attended by rheumatologists through phone calls. Analytical blood test and X-rays was performed in 89 and 77 patients respectively.

Regarding persistent symptoms after discharge (table 2), 68.57% of patients had at least one symptom, and 26.84% had more than three. In those with symptoms, the most frequent reported was dyspnea (52.77%), fatigue (37.5%), chest pain (25%), cough and diarrhea/vomiting (19.4%). All had an average duration of more than one month, with diarrhoea and fever having the shortest duration. The longest reported duration were over three months for alopecia and physical deconditioning.

**Table 2.** Symptoms and consequences in RMD patients discharged after COVID-19. Symptoms median duration report the lag time from discharge until the last date the event was reported. Consequences median duration report the lag time from discharge until the development report.

|  | n (%) | Median (IQR), days |
| --- | --- | --- |
| **Self-reported symptoms (n=105)** | 72 (68.57) |  |
| Dyspnea | 38 (36.2) | 68.5(23-97.5) |
| Fatigue | 27 (25.71) | 38.5(14-83) |
| Chest pain | 18 (17.14) | 30(14-150) |
| Cough | 14 (13.33) | 27(18-99) |
| Diarrhoea/vomiting | 14 (13.33) | 31(15-55) |
| Fever | 11 (10.48) | 29(22-48) |
| Anosmia/ Dysgeusia | 9 (8.57) | 92(25-134) |
| Anxiety/Depression | 8(7.62) | 56.5(22.5-73) |
| Dermatological manifestations | 7 (6.67) | 37(27-68) |
| Joint pain | 6 (5.71) | 55(20-68) |
| Visual/ auditory alterations | 5 (4.76) | 53(18-112) |
| Headache | 5 (4.76) | 28(12.5-85.5) |
| Physical deconditioning | 4(3.81) | 111(45.5-138.5) |
| Hair loss | 2(1.90) | 83.5(83-84) |
| Cognitive dysfunction | 1 (0.95) | 20 |
| **Consequences (n=105)** | 31 (29.5) |  |
| Death | 2(1.90) | 13.2(8-19) |
| Analytical abnormalities (n=89) | 16 (17.97) | 45.5 (21-81) |
| Lung damage | 12 (10.47) | 44(24-77.5) |
| Oxygen necessities | 6 (5.7) | 20.5(7-47) |
| Thrombotic event | 1(0.95) | 42 |
| Optic neuritis | 1(0.95) | 27 |
| Rehabilitation needed (physical/respiratory) | 5(4.76) | 54(43-88) |

**Table 3.** Multivariable analysis. Risk factors for persistent symptoms after COVID-19 in RMD. Independent variable RMD group.

| Variable | OR | 95% CI | p |
| --- | --- | --- | --- |
| **Gender, women** | 3.53 | 1.24-10.08 | 0.01 |
| **Age, years** |  |  |  |
| <50-60 | 1 | - | - |
| 60-75 | 0.11 | 0.02-0.48 | 0.004 |
| >75 | 0.09 | 0.02-0.4 | 0.002 |
| **ARD vs NARD** | 0.43 | 0.15-1.23 | 0.12 |
| **Baseline number of comorbidities \*** | 1.90 | 1.09-3.31 | 0.023 |
| **Lymphopenia\*\*** | 3.17 | 1.07-9.4 | 0.038 |
| **Pneumonia\*\*\*** | 10.1 | 1.5-66.01 | 0.017 |

\* (ILD/COPD);\*\*Lymphopenia: less than 800 lymphocytes during the admission; \*\*\*Presence of pneumonia during admission. \*\*\* Defined as the number of comorbidities prior COVID admission. They include the follows: hypertension, heart disease, ischemic vascular disease, diabetes mellitus, venous thrombosis/lung embolism, chronic kidney disease, liver disease, lung disease (ILD/COPD)

We did not find statistical significance among ARD (68.5%) and NARD (68.6%) patients with persistent symptoms. In ARD, the mean number of symptoms was 1.48(1.46), and for NARD was 1.64(1.65), without significant difference. As we show in figure 2, type of persistent symptoms were very similar among ARD and NARD patients, with slight differences in chest pain (p=0.3), gastrointestinal symptoms (p=0.3), anosmia/ dysgeusia (p=0.3) and visual-auditory alterations (p=0.2).

**Figure 2.**
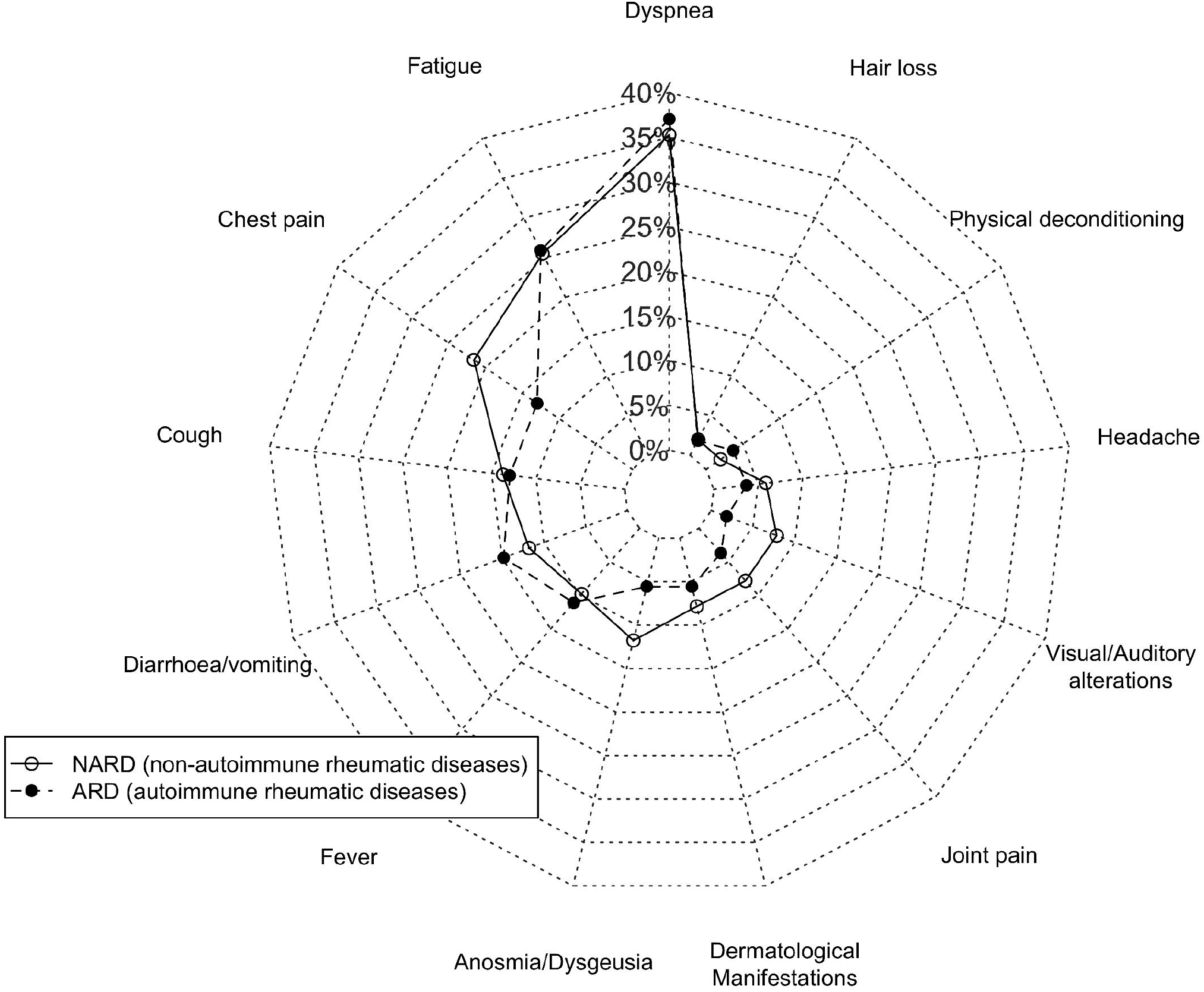
Persistent symptoms after Covid-19 (ARD and NARD patients)

During the follow-up post discharge, 29 patients returned 31 times to the emergency room and 14(50%) required admission, being 78.6% of them (n=11) related to Covid-19 (mean and median lag time: 31.6(28.5); 19 (p25-75: 8-53) days). The latest were caused by lung problems, thrombotic problems, kidney failure, hepatitis, and cutaneous anaphylaxis. There was also one readmission due to Covid-19 reinfection in ARD patient. We found other COVID-19 reinfection after five months that did not require admission (NARD). Interestingly, ARD patients (16.7%; 33% of them due to lung or renal infections) had a higher percentage of covid-19 related readmissions than NARD (3.9% and none infections), almost achieving statistical significance (p=0.053).

Regarding the 77 patients with radiologic imaging during the follow-up, 75 of them improved step by step overtime (mean and median lag time: 61(29); 54 (41-74) days), and 20 of them (26.6%) achieved completely recovery in radiologic image during the follow-up period. Two patients got worse, one developed a lung cancer and the other a Pneumothorax. Six patients required oxygen support as a consequence of lung damage after Covid-19, being similar among RMD groups.

We considered lung damage as consequence evaluating radiographic image, pulmonary tests function, clinical evolution and oxygen support after discharge. From all, 12 patients (11.4%) were considered to have lung damage over time (table 2; figure 3): 2 with bacterial infection that required a new hospital admission (mean time: 29 (20.5) days), 1 lung cancer (after 74 days), 3 with previous COPD disease worsening their baseline function (one of them developed an atelectasis), (mean time: 43.5 (10.5) days), and 6 interstitial residual interstitial pneumonitis (mean time: 89.2 (53) days). From the latest, one of them developed recurrent pneumothorax that required hospital admission at least three times during the study period (the first after 8 days of discharge), another had previous interstitial lung disease worsening after the acute infection. Although ARD patients had more percentage of lung damage, these differences did not achieve statistical significance (p=0.7).

**Figure 3.**
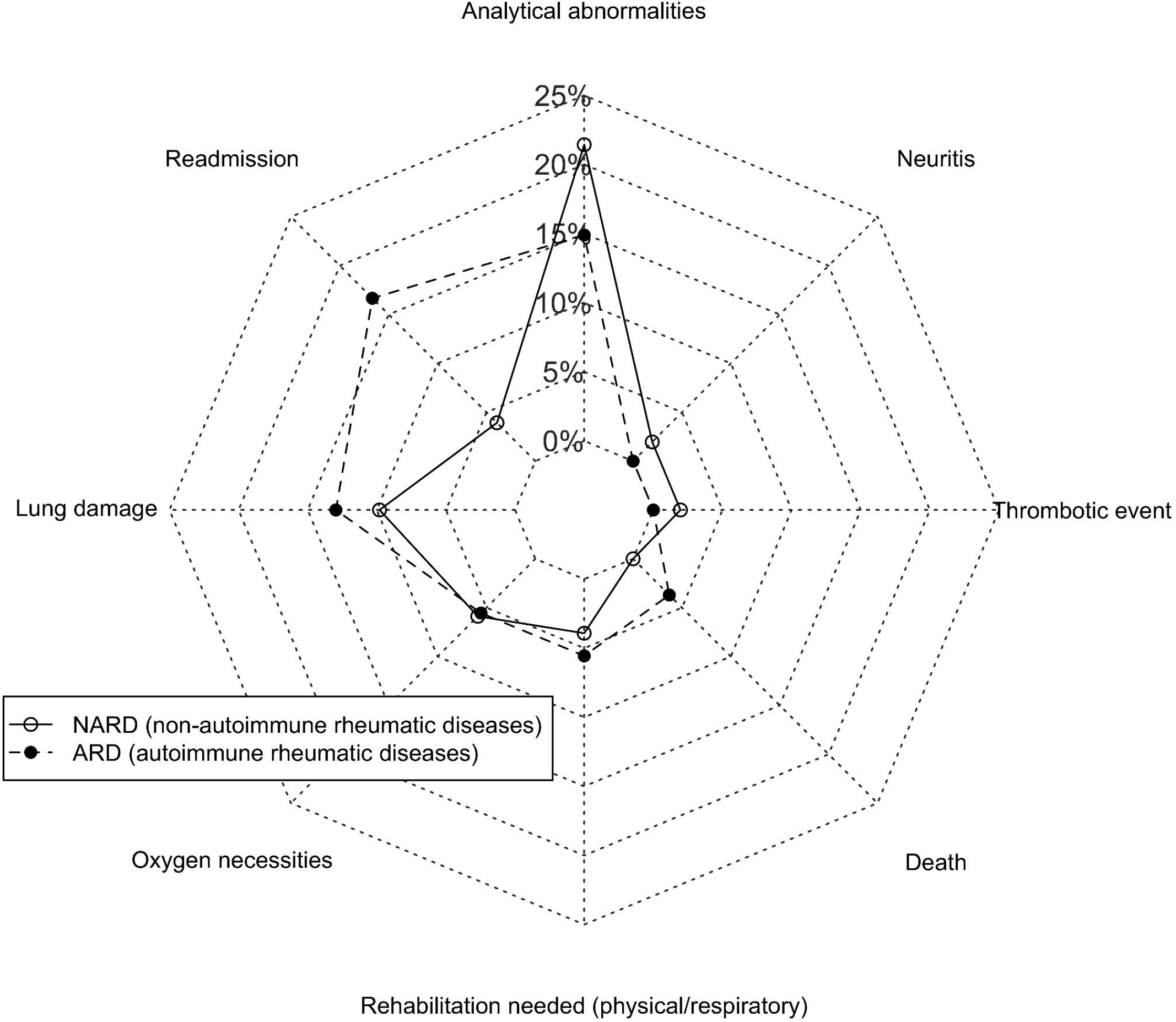
Health Consequences after Covid-19 (ARD and NARD patients)

From those patients that had blood tests performed, 18% of them developed abnormalities that were not present prior to admission: 9 (10%) with lymphopenia lower than 800, 4 with elevated liver transaminases, 5 with renal failure and 2 with thrombopenia. 14% of ARD had blood test alterations compared to 21% of NARD, without statistical significance (p=0.4). Specifically for lymphopenia ARD had 11% whereas NARD had 5.9% (p=0.4). Two patients died during the follow-up. They had ARD and their median age was 87.5 (84-91) years. One of them died at home eight days after discharge and had relevant comorbidity including previous lung disease. The other patient died during a hospital readmission 87 days after the discharge, due to a urinary sepsis.

Regarding other consequences, one patient developed a central retinal vein occlusion and the remaining patient developed optic neuritis. Both of them were NARD patients (figure 3). Finally, a total of five patients required intensive rehabilitation, 4 of them with associated physical deconditioning symptoms. Encompassing all, 31 patients had at least one consequence resulting from the Covid-19 disease. The prevalence was similar among RMD groups (ARD: 29.6%: NARD: 29.4%; p=0.9).

ARD had lower risk of persistent symptoms compared to NARD, but without statistical significance in the univariate (OR: 0.99, [0.43-2.27]) and multivariate analysis (table 4). In the latest, prior comorbidities, female and younger patients had more probability of post-discharged symptoms. Other factors such as lymphopenia or pneumonia during admission also achieved statistical significance in the final model.

Concerning the influence of RMD group on consequences, we did not find differences between ARD compared to NARD neither in the univariate or multivariate analysis (OR: 1.85 [0.67-5.14]; p=0.23). We did not find statistical difference between age groups, neither baseline comorbidity, whereas female compared to men [HR 0.28 (0.10–0.77), p=0.014] had less probability of consequences and the presence of Covid-19 complications during the admission [HR 9.54 (3.16– 28.8), p= 0.001] increased the risk.

## Discussion

Our study has identified that a higher number of RMD patients after Covid-19 admission still have persistent symptoms, even 6 months after discharge, especially fatigue, dyspnea, anosmia and chest pain. This study also evaluates health consequences and highlights that lung damage seems to be the most frequent in these patients. ARD compared to NARD patients were similar in persistent symptoms or consequences, although ARD patients might have a greater number of readmissions.

Available data regarding persistent symptoms after Covid-19 are scarce and heterogeneous, but all show it is a frequent issue [(8)(11)]. In our study near 70% reported persistence of at least one symptom.

Almost 40% of the study patients reported dyspnea with an average duration of more than two months, being the most frequent persistent symptom, as in other published studies, [(11)(13)(19)]. Our results about persistent fatigue, cough or gastrointestinal symptoms are also in line with published data[(11)(13)(20)].

For persistent joint and muscular pain, other studies has been reported about 10 to 27% at 4-8 weeks after infection [(13)(11)(19)]. Our follow-up is similar, but the percentages found are lower, probably due to most of RDM patients had it previously and do not report it as a consequence of Covid-19.

In some other symptoms, such as anosmia, headache, physical deconditioning or anxiety or depression, our study reported lower percentages compared to other studies [(13)(11) (19)(21)(22)(23)(24)]. Nevertheless, we have to take into account that data in our study are retrospectively collected from medical records and patients were not specifically asked for them. Maybe those symptoms were not reflected in the clinical history because they were considered less relevant than others for the physician or the patient, or maybe they were symptoms that the patient did not report because they had not bothered the patient enough. Covid-19 pneumonia still had residual interstitial abnormalities in some patients over three months, and others worsen their COPD two months after suggesting that in a group of discharged patients, lung damage may persist. Other studies have also reported a significant percentage of patients with abnormal results in respiratory function and imaging studies afterCovid-19 acute infection [(20)(25)(26)]. Almost 12% of our RMD patients developed lung disease or worsen their lung damage after discharge, being 39% of the total health consequences found in our sample.

Regarding thrombotic events and in accordance with our findings, other studies reported a prevalence of 0.5-2.5%[(27)(28)]. We reported one ocular thromboembolic event as it has been described elsewhere (29).

During the follow-up, 27.6% of the patients returned to emergency room due to a health problem. From them, 50% required a new hospital admission in the first 30 days after having overcome Covid-19. In the study of Donelly et al the percent of readmission was somewhat higher(30).

Four discharged patients received chest physiotherapy. Although this data fits with the patients who manifested deconditioning, if we compare it with the persistence of dyspnea, it seems a low number. In this sense, more attention should be given to the management of post-Covid-19. Patients under oxygen support would benefit from low intensity exercise, according to the recommendations of Stanford Hall consensus statement for post-Covid-19 rehabilitation (31).

Musculoskeletal specific impairments related to post-Covid-19 are not well known yet. In patients who have been unconditioned for a long time, their muscles, joints may worsen without physical therapy intervention and even more in those with previous RMD. Aerobic exercise and adapted physical activity interventions are considered the main beneficial interventions (32). In those hospitalized patients with comorbid conditions, long admission and in those requiring intensive care, rehabilitation would be mandatory.

Fortunately, despite excess patient mortality during hospital admission at the peak of the pandemic, 1.9% of the patients died as a consequence of Covid-19 in the first month after hospital discharge but no one else until the end of study.

Concerning the influence of RMD group on persistent symptoms, the risk for ARD was lower but without statistical significance. This slight difference could related to the tight control of patients with ARD, including a strict management of their comorbidities, especially cardiovascular risk factors with statins [(33)(34)], or osteoporosis, treated with vitamin D [(35)], but also due to DMARDs that might have a beneficial effect avoiding complications associated with Covid-19. Another interesting finding is that female, younger patients, presence of lymphopenia or pneumonia during admission were factors independently associated to persistent symptoms. In accordance with the results of Cook et al (36), our study also reveal that baseline comorbidities can increase the risk of persistent symptoms.

When the consequences are analyzed, we did not find differences between ARD and NARD patients, although being male and Covid-19-related complications during admission increased the risk. However, the number of readmissions was higher in ARD patients. probably due to their risk of infections and complications associated with their immunosuppression. Post-infection care for Covid-19 survivors add an additional burden to the healthcare system, even strained by the challenges of the pandemic. The follow-up of these patients has been close after hospitalization, especially in ARD patients, with an important role played by the rheumatology nurses.

Our study has several limitations. First, it is a retrospective observational study, and we cannot assure that everything has been found and described. It is cross-sectional, not being able to capture how the problems evolve over time. In this sense, longitudinal studies specifically designed to assess the post Covid-19 evolution over time would be necessary to better understand the progression of symptoms and consequences after Covid-19.

Notwithstanding these limitations, we believe is necessary acquiring “real-life” data after the acute phase of Covid-19 in RMD. In this sense, our study give us a general picture of what is going on in RMD patients spanning a period of up to 7 months. Although they are preliminary findings, this study provides rheumatologists and ARD patients some reassurance since their evolution does not seem worse that other post-Covid-19 patient, with the exception of higher percentage of readmissions found in ARD. In this sense, closer monitoring would be advisable. This study also give as a clue of which factors could influence in the post-Covid-19 phase.

To conclude, we provide a report of symptoms and consequences, after a hospitalized episode of Covid-19 in RMD patients. This study is a starting point to advance in knowledge. Longer follow-up studies in a larger RMD cohorts are necessary to understand the full spectrum of health consequences from Covid-19 and to design future care plans.

## Data Availability

Data are available upon reasonable request. The data will be shared if there is a reasonable request for it.

## Contributors

LL and LA conceived and designed the study. LL, LA IP, LLP, JIC collected data. LA, AM and LL performed the data analysis and interpreted the data. All of the authors were involved in the drafting and/or revising of the manuscript.

## Disclaimer

Funders had no impact on the design or interpretation of the study or its results.

## Ethics approval

The study was approved by the Hospital Clínico San Carlos institutional ethics committee (approval number 20/268-E-BS). This study was conducted according to the principles of the Declaration of Helsinki.

## Patient and Public Involvement

Patients or the public were not involved in the design, or conduct, or reporting, or dissemination plans of our research.

